# A Demographic Look at Cancer Treatment Behaviors during the COVID-19 Pandemic

**DOI:** 10.64898/2026.03.24.26349229

**Authors:** Jonathan Acosta Morales

## Abstract

**Background:** While numerous studies have explored the relationship between COVID-19 and cancer, few have specifically examined the significant impact of the pandemic on cancer patients, particularly concerning their treatments and appointments.

**Objectives:** This study aims to investigate cancer treatment behaviors during the COVID-19 pandemic.

**Methods:** This retrospective quantitative study utilized data from the Centers for Disease Control and Prevention’s National Health Interview Survey of 2020. The inclusion criteria were as follows: studies conducted within the United States; patients diagnosed with COVID-19 since the pandemic began; patients diagnosed with cancer within the United States; patients undergoing cancer treatment or in remission since the start of the pandemic; patients who experienced a change, delay, or cancellation of treatment due to the COVID-19 pandemic; patients who experienced a change or delay in cancer care due to the COVID-19 pandemic; patients with a weakened immune system due to prescriptions; and patients who took prescription medication within the past 12 months. The variables were analyzed against population characteristics, including age, race, gender, cancer type, and COVID-19 status. Python Jupyter Notebook (packaged by Anaconda Navigator in R Studio, version 6.4.8), Microsoft Excel for data cleaning and assessment, and SPSS were used for statistical analyses.

**Results:** Chi-Square Analysis (p<.05) revealed significant associations between cancer treatment and gender (p=0.009), other cancer treatments and age (p<.001) and education (p<.001), changes in other cancer treatments and gender (p=0.045), race (p<.001), age (p<.001), and education (p=.013), and prescribed medication and gender (p=.009), family income (p<.001), and age (p<.001).

**Conclusion:** The COVID-19 pandemic has significantly impacted cancer care in the U.S., affecting the delivery of treatments. Additional government funding is necessary to help medical facilities develop programs for off-site treatment delivery, to better prepare for future pandemics, and avoid repeating past challenges.

## 1 Introduction

The novel COVID-19 virus swept the United States and disproportionately impacted the lives of the citizens, especially cancer patients, around the nation. The cancer patients affected include those undergoing active treatment, those who have completed treatment, and patients in remission. ^1,2^ The COVID-19 virus is a threat to public health and societal well-being. The U.S. Centers for Disease Control and Prevention (CDC) reports that COVID-19 mortality rates were equivalent to 190/100,000 of the African Americans, Native Americans, and Latino ethnic groups.^4^ Unfortunately, the disparities in COVID-19 experienced by minorities are consonant with similar comorbidities, such as obesity, diabetes, asthma, chronic obstructive pulmonary disease, hypertension, cardiovascular disease, and kidney disease, as distinguished in cancer. ^5,6,7,8,9.^

Evidence shows that patients may experience clinical symptoms similar to the flu and common colds, such as dry cough, fever, diarrhea, vomiting, and myalgia, while patients with numerous comorbidities may experience severe infections, such as acute respiratory distress syndrome and kidney damage^.3,10,11^ Due to the immunosuppressive nature of some cancers, sudden shifts in health care services and management of cancer patients in the U.S. changed.^12^ Since the onset of the COVID-19 Pandemic, cancer patients have experienced a disruption in regulatory access to health care and cancer treatment. Leading to gaps in outpatient cancer clinical visits, cancer screening rates, and enrollment in clinical trials, all while experiencing a decrease in emotional and mental health.^13^ Although there have been many studies about the relation of COVID-19 with cancer, there are limited studies that have examined the significant impact of the COVID-19 Pandemic on cancer patients and the effects that social distancing regulations mandated by the government had on prolonged patient treatments and appointments.

The purpose of this study is to investigate the significant differences among selected demographic factors (age, race, and gender) in the impact of the COVID-19 pandemic on the healthcare services provided to cancer patients in the U. S. The overarching hypothesis of this study is that the COVID-19 pandemic’s impact on the quality of healthcare services will disproportionately affect cancer patients, with variations observed across different demographic factors (age, race, and gender) in the U.S.

Using a quantitative retrospective approach, pulling pre-existing data from the CDC’s National Health Interview Survey (NHIS) of 2020.^14^ The data was analyzed to pinpoint the most significant impact of the COVID-19 pandemic on cancer patients’ health care services and management of care.

## 2 Research Method

In this quantitative retrospective study, data from the CDC’s National Health Interview Survey of 2020^14^ was utilized, encompassing 31,568 participants aged 18 years and older across the United States. The CDC conducted interviews via telephone from March 2020 through June 2020 and both in-person and by telephone from July 2020 to December 2020. Initially, the sample size for this study included 4,130 cancer patients who responded to the NHIS survey. However, 122 participants were excluded due to missing information in any category, reducing the final sample size to 4,008 participants.

Participants were included if they were dually diagnosed with COVID-19 and cancer. The study analyzed various characteristics such as age, race, gender, cancer diagnosis, COVID-19 diagnosis, household members, household region, multi-race status, educational level, family income, interview month, health status, cancer and COVID-19 treatment, changes in cancer treatment, changes in other cancer care, and prescription medication. Those who did not meet these criteria were excluded. The variables utilized from the NHIS survey included “CVDDIAG_A”, “CANEV_A”,”CANCOVTREA_A”, “CANCOVCHG_A”, “CANCOVOTH_A”,”CANCOVCARE_A”,”MEDRXTRT _A”, “RX12M_A”, “SEX_A”, and “RACEALLP_A”.

The primary independent variable in this study is whether participants were currently infected with COVID-19, immunosuppressed due to their cancer condition, or dually affected by both. The dependent variable focuses on the changes, delays, or cancellations of cancer treatment as a result of the COVID-19 pandemic or active infection. The hypothesis posits that the COVID-19 pandemic has a more adverse impact on the quality of healthcare services for cancer patients, with this effect varying based on demographic factors (age, race, and gender) in the U.S. This study aims to elucidate the differential impact of the pandemic on cancer care, highlighting the disparities influenced by these demographic variables.

Data quality was ensured by checking for missing data, removing outliers, and transforming variables to avoid biases. High healthcare service levels were defined as normal treatment schedules, while low healthcare service levels were defined as delayed and/or canceled treatments due to the pandemic. Descriptive statistics were performed on frequency, mean, and percentile. An independent t-test was conducted using Chi-Square (X^2^) analysis to determine if there were differences in the variables of interest. Statistical analyses were conducted using Excel, Python Jupyter Notebook (version 6.4.8), and SPSS.

## 3 Result

### In Table 1: Characteristics and Frequency

The analysis of participant characteristics reveals several notable trends and distributions across different demographic factors. The gender distribution shows a higher proportion of females (57.8%) compared to males (42.1%). The vast majority of participants identified as White (91.5%), with smaller percentages identifying as Black/African American (5.9%), Asian only (1.2%), American Indian or Alaska Native (AIAN) only (0.4%), and those identifying as AIAN and any other group (0.9%). Only 2 participants identified with other single and multiple races. Health status varied among participants, with the highest percentages reporting their health as either very good (31.4%) or good (32.1%), and smaller proportions rating their health as excellent (12.6%), fair (16.7%), or poor (7.1%). Most participants lived in two-person households (88.7%), with significantly fewer in households of other sizes: one-person (3.1%), three-person (5.7%), four-person (1.2%), five-person (0.3%), and six or more persons (0.9%). Family income levels were fairly evenly distributed, with the largest groups earning either $0-$34,999 (28.7%) or $100,000+ (27.1%), and the remaining participants reporting incomes of $35,000-$49,999 (14.0%), $50,000-$74,999 (17.6%), and $75,000-$99,999 (12.6%). Age distribution showed that participants predominantly fell into the older age brackets, with 52.9% aged 56-75 years and 33.1% aged 75-95 years. A smaller portion of participants were aged 36-55 years (12.4%) or 18-35 years (1.6%). Education levels among participants were varied, with the largest group having some college or an associate’s degree (29.8%), followed by those with a high school diploma (24.7%). Smaller groups included those with a bachelor’s degree (21.5%), a master’s degree or above (18.3%), and less than a high school diploma (5.8%).

***Table 2: Chi-Square Analysis*** presents a detailed analysis of the impact of the COVID-19 pandemic on cancer treatment across various demographic factors, highlighting several significant findings. There were significant gender differences, with females (94.0%) being more likely to not receive cancer treatment since the pandemic started compared to males (91.8%), and also more likely to take prescription medication (91.7% vs. 89.2%), both with a p-value of 0.009. Females (4.2%) were also more likely to experience changes in secondary cancer care than males (3.0%), with a p-value of 0.045. Racial disparities were notable, particularly for Asian participants, who had a significantly higher rate of changes in secondary cancer care (22.2%) since the pandemic began, with a p-value of <0.001. Prescription medication use varied significantly with family income, with those earning $50,000-$74,999 (16.2%) and $75,000-$99,999 (11.3%) less likely to use prescription medications, p-value <0.001. Age also influenced treatment, with older participants (56-75 years at 48.0% and 75-95 years at 31.4%) significantly more likely to use prescription medications and less likely to receive secondary cancer care, both with p-values of <0.001. Additionally, participants aged 36-55 had a higher rate of changes in secondary cancer care (6.2%), p-value <0.001. Educational attainment also showed significant differences; participants with some college or an associate’s degree (26.4%) were more likely to take prescription medications, and those with a master’s degree or above (5.7%) were more likely to experience changes in secondary cancer care, with p-values of <0.001 and 0.013, respectively. These findings underscore the significant disparities in cancer treatment and care during the pandemic across different demographic group.

**Table 1:**
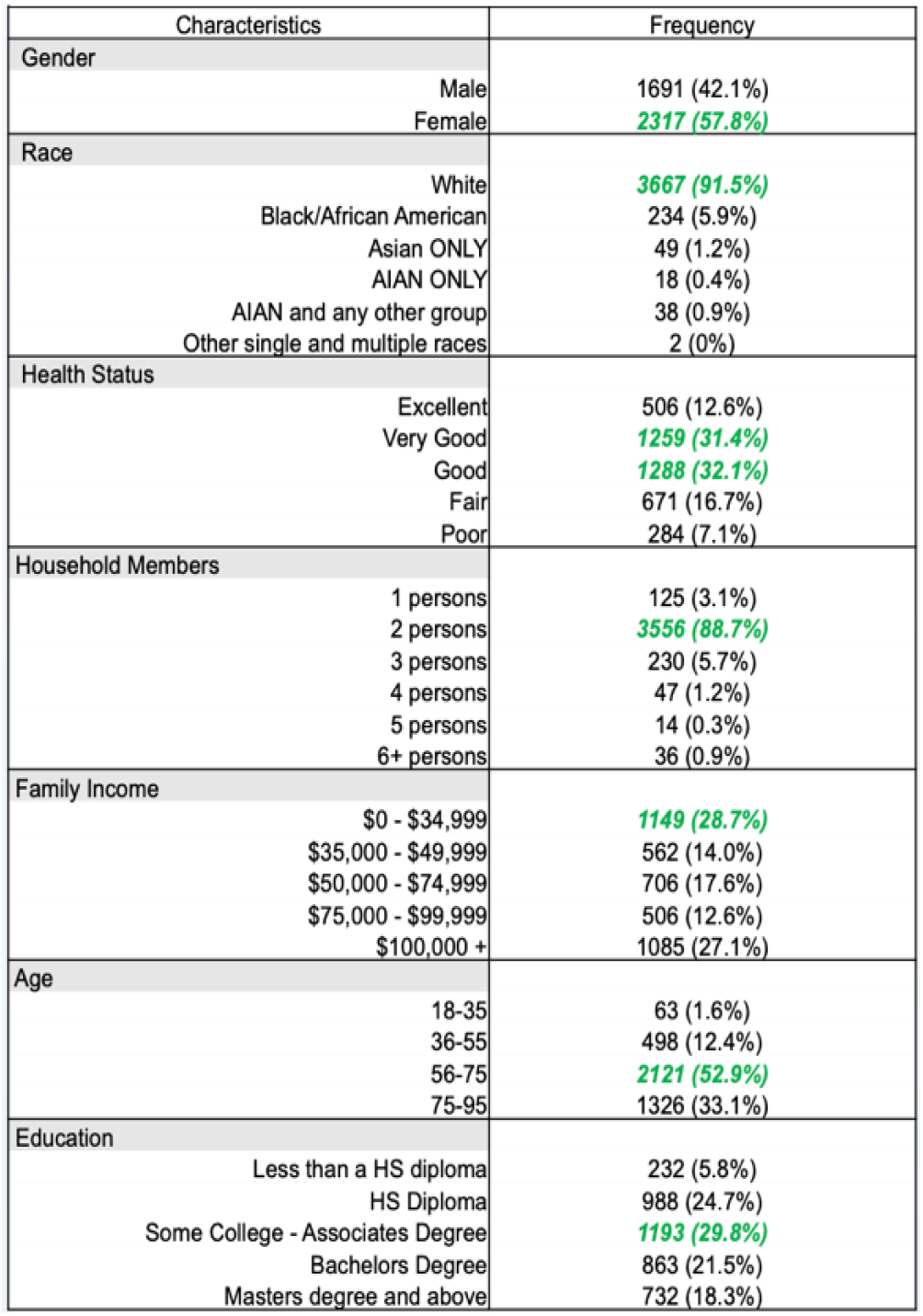
Characteristics and Frequency:

**Table 2:**
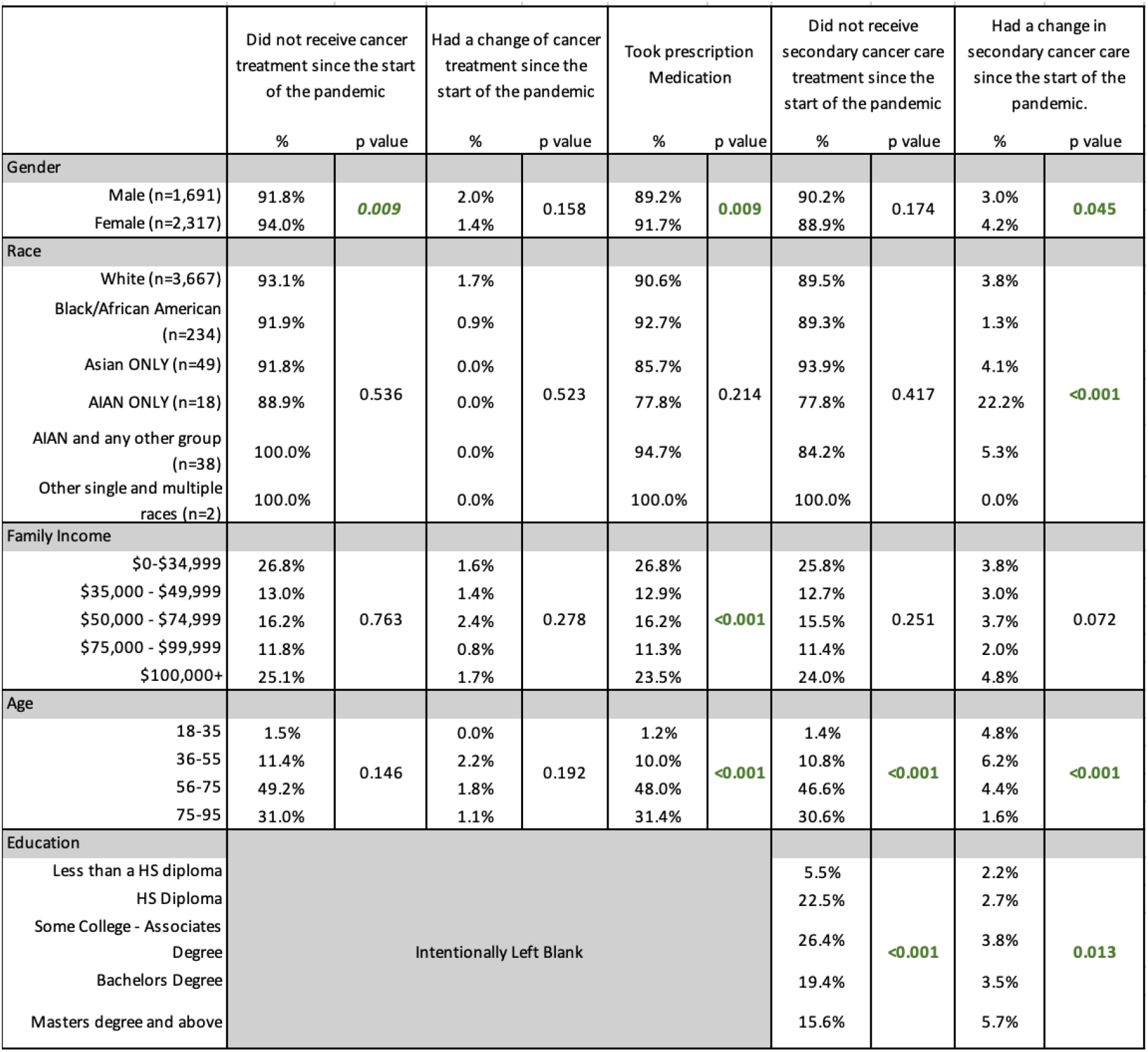
Chi-Square Analysis (p<.05) revealed significant associations between cancer treatment and gender (p=0.009), other cancer treatments and age (p<.001) and education (p<.001), changes in other cancer treatments and gender (p=0.045).

**Table 3:** Chi-Square Analysis.

## 4 Discussion

The Chi-Square Analysis indicated a significant association between cancer treatment and gender (p=0.009). Female patients were more likely to experience changes, delays, or cancellations in their cancer treatment compared to male patients. This finding aligns with existing literature that suggests women may face more barriers to accessing healthcare^15^, possibly due to caregiving responsibilities and other social factors that influence their ability to seek timely treatment.^2^

Age was a significant factor in the disruption of other cancer treatments (p<0.001). Older patients (aged 65 and above) were particularly affected, experiencing more frequent interruptions in their treatment schedules. This may be due to heightened precautions taken to protect older^16^, more vulnerable populations from COVID-19, as well as the increased risk of severe outcomes if they contracted the virus. ^10^

Educational level also showed a significant association with changes in other cancer treatments (p<0.001). Patients with lower educational attainment were more likely to experience disruptions in their treatment.^17^ This could be attributed to a lack of access to information about alternative treatment options or lower health literacy, which may hinder their ability to navigate the healthcare system effectively during the pandemic.^6^

Gender (p=0.045), race (p<0.001), age (p<0.001), and education (p=0.013) all showed significant associations with changes in other cancer treatments. Minority patients, including African American, Native American, and Latino groups, experienced more significant treatment disruptions compared to White patients. This disparity reflects broader systemic inequities in healthcare access and quality, which have been exacerbated by the pandemic^.5,18^

The analysis revealed significant associations between prescribed medication and gender (p=0.009), family income (p<0.001), and age (p<0.001). Female patients, those with lower family income, and older patients were more likely to report disruptions in their prescribed medication regimes.^19^ These findings suggest that economic factors and social determinants of health play a critical role in the ability of patients to maintain their medication schedules during a crisis.^13^

The study further analyzed various variables such as household members, household region, multi-race, educational level, family income, interview month, health status, cancer and COVID-19 treatment, changes in cancer treatment, change in other cancer care, and prescription medication. This comprehensive analysis provided a detailed understanding of how the pandemic impacted different aspects of cancer care across various demographics.

For instance, patients from lower-income households or those residing in rural areas reported more significant challenges in accessing cancer care. The shift from in-person to telephone interviews by the CDC also highlighted disparities in communication and access to healthcare services, particularly for those without reliable telephone or internet access (NHIS, 2020).^14^

The data also indicated that patients who were already experiencing multiple comorbidities faced compounded difficulties during the pandemic. These patients were more likely to report severe disruptions in their cancer care, underscoring the need for targeted interventions to support high-risk groups during public health crises.^3^

The statistical significance of these findings supports the hypothesis that the COVID-19 pandemic had a more adverse impact on cancer patients’ treatment behaviors, influenced by age, race, gender, and socioeconomic status.

## 5 Conclusion

The COVID-19 pandemic has significantly disrupted cancer care in the United States, revealing and exacerbating existing healthcare disparities.^20^ Our study found that demographic factors played a critical role in the extent of these disruptions. Older adults, women, minority groups, and those with lower educational attainment or income were particularly affected.

These findings highlight the urgent need for policy interventions and additional funding to support equitable healthcare delivery, especially during public health emergencies. Programs that facilitate off-site or home-based cancer care could mitigate the impact of such disruptions in the future.^21^ Furthermore, addressing the mental health needs of cancer patients through integrated care models is essential to improve their overall well-being.

To enhance the validity and reliability of future studies, there is a need for a more balanced female to male ratio, more evenly distributed races among participants, and a broader representation of age groups. A more diverse and representative sample will provide a more comprehensive understanding of the pandemic’s impact on various demographic groups and contribute to more effective and inclusive healthcare policies and interventions.

## Data Availability

All data analyzed in this study are from the publicly available 2020 National Health Interview Survey (NHIS), which can be accessed through the CDC/NCHS website and IPUMS NHIS.

https://www.cdc.gov/nchs/nhis/documentation/2020-nhis.html

## 6 Future Direction

Future research must include longitudinal studies to track the long-term impact of the COVID-19 pandemic on cancer care outcomes. It’s crucial to develop and implement sustainable and robust telehealth care protocols capable of withstanding public health crises. Policy advocacy is essential to secure increased funding and resource allocation for cancer care during emergencies, which will help minimize the adverse effects on vulnerable populations. Furthermore, additional government funding is needed to support medical facilities in creating off-site treatment programs, ensuring better preparedness for future pandemics and avoiding past challenges.

## 7 Acknowledgements

The author extends their gratitude to Dr. Adiebonye Jumbo, PhD, ITIL, for her mentorship, to Tina Adjei-Bosompem, MPH, for assistance with biostatistics and Chi-Square analysis, and to Haley Rippy for presentation edits and support during the SPRINTER Symposium. They also thank Maimouna Ndiaye, MPH, for her insightful lectures on health disparities. Appreciation is given to SUNY Downstate Health Sciences University for their collaboration with CUNY Hunter College. The author is grateful to the SPRINTER program for their support and to the Brooklyn Health Disparities Center and the TRANSPORT/NIH grant for funding this research.

## 8 Financial Disclaimer

This research project was funded by the National Institute of Health Grant #: 5S21MD012474-02, received by SUNY Downstate University and the Brooklyn Health Disparities Center through the Translational Program of Health Disparities Research Training (TRANSPORT).

